# Multi-task analysis of gene expression data on cancer public datasets

**DOI:** 10.1101/2023.09.27.23296213

**Authors:** Yasmmin Martins

## Abstract

**Background:** There is an availability of omics and often multi-omics cancer datasets on public databases such as Gene Expression Omnibus (GEO), International Cancer Genome Consortium and The Cancer Genome Atlas Program. Most of these databases provide at least the gene expression data for the samples contained in the project. Multi-omics has been an advantageous strategy to leverage personalized medicine, but few works explore strategies to extract knowledge relying only on gene expression level for decisions on tasks such as disease outcome prediction and drug response simulation. The models and information acquired on projects based only on expression data could provide decision making background for future projects that have other level of omics data such as DNA methylation or miRNAs.

**Results:** We extended previous methodologies to predict disease outcome from the combination of protein interaction networks and gene expression profiling by proposing an automated pipeline to perform the graph feature encoding and further patient networks outcome classification derived from RNA-Seq. We integrated biological networks from protein interactions and gene expression profiling to assess patient specificity combining the treatment/control ratio with the patient normalized counts of the deferentially expressed genes. We also tackled the disease outcome prediction from the gene set enrichment perspective, combining gene expression with pathway gene sets information as features source for this task. We also explored the drug response outcome perspective of the cancer disease still evaluating the relationship among gene expression profiling with single sample gene set enrichment analysis (ssGSEA), proposing a workflow to perform drug response screening according to the patient enriched pathways.

**Conclusion:** We showed the importance of the patient network modeling for the clinical task of disease outcome prediction using graph kernel matrices strategy and showed how ssGSEA improved the prediction only using transcriptomic data combined with pathway scores. We also demonstrated a detailed screening analysis showing the impact of pathway-based gene sets and normalization types for the drug response simulation. We deployed two fully automatized Screening workflows following the FAIR principles for the disease outcome prediction and drug response simulation tasks.

**Availability:** The ScreenDOP code is available at https://github.com/yascoma/screendop while the DReCaS is available at https://github.com/YasCoMa/caliscoma_pipeline/

## 1 Introduction

The wide availability of public biological datasets, specifically related to the omics data analysis lead to a leverage of bioinformatic applications taking advantage of these pieces of curated data for deep and machine learning applications in biomedical research problems. Some of these applications may be drug discovery and repurposing [1], disease prognosis [2] and diagnosis [3] prediction, biomarker discovery [4] and precision medicine [5]. The most popular type of omic dataset is the transcriptomic data, there are several curated and documented experiments in the Gene Expression Omnibus (GEO) database using either Microarray or RNASeq techniques [6]. Specifically for cancer disease, there are initiatives such as The Cance Genome Atlas^1^ (TCGA) and the International Cancer Genome Consortium^2^ (ICGC) that provides several projects with samples metadata and at least one type of raw data is publicly available, and among this data often a project contains a multi-omic perspective, but the majority offer gene expression data.

The transcriptomic data, depending on the way the experiment was design, is a powerful source of information that is applied for features processing in machine learning applications. Some of the biomedical tasks in which this type of data figures are the disease outcome prediction [7–9] and drug response simulation [10]. Recent methodologies for both task have been taking advantage of multi-omic information, but in order to acquire knowledge from past experiments that only provide transcriptomic data, the multi-omic strategies could miss information to perform correctly when transferring learning. Few methods regarding precision oncology [11], for both tasks, provide their methods in a way that allows reproducibility and handles all the process since data preparation for the method application to the post processing analysis and output exportation, following the FAIR (Findable, Accessible, Interoperable and Reusable) principles [12].

In this paper, in line with the FAIR principles and allowing reproducible research, we present two workflows (ScreenDOP – Screening or Disease Outcome Prediction and DReCaS – Drug response calibration simulation) respectively for disease outcome prediction and drug response calibration simulation tasks, that allows the execution of multiple experiment screening in a fully automated ans transparent manner. We aimed at providing an end-to-end data process from acquirement and treatment to the results exportation and analysis, allowing the hypotheses tracking and performance explainability.

The ScreenDOP was designed two offer two strategies to extract the numerical features for posterior application in a classifier, one of the strategies proposes the combination of gene expression with protein interactions and using kernel functions form the features with the samples graph similarity matrix. We extended and generalized the application of the first strategy previously used on [13] to RNA-Seq with multiple experimental design matrices and allowed the modulation of the nodes/edges attributes that are used by the kernel functions. The second strategy combines the gene expression data with gene set enrichment analysis (GSEA) to extract pathway scores. This second method aims at reducing features dimensionality and consequently increasing computational efficiency in the workflow execution.

The DReCaS workflow was projected to automatize the data preparation and allow the method proposed by [14], which calibrates patient-specific drug responses, be automatic and flexible to a diversity of execution scenarios (changes of normalization types and pathway gene set libraries). We also extended its methodology to allow the transfer learning mode from a complete cancer dataset with healthy and disease samples to the disease ones of of another dataset belonging to the same type of cancer. Two other new features were enabled in DReCaS that are the optimization of the weights used to calibrate the samples scoring matrix, and the possibility to evaluate a batch of individual drug or a list of specific drug combinations, forming a report of drug/combination prioritization and ranking.

https://www.sciencedirect.com/science/article/pii/S0079610722000803

https://www.mdpi.com/2073-4425/10/3/238

In line with the FAIR principles and allowing reproducible research on drug discovery experiments, we proposed a drug response simulation workflow inspired on a previous work with a flexible data input handling, parameter optimizationa and fully automatized from data preprocessing to drug test screening. https://www.nature.com/articles/s41597-019-0174-7

## 2 Methods

We proposed a framework (ScreenDOP – Screening for Disease Outcome Prediction) that offers two strategies to extract knowledge from the expression data provided by the GEO database from NCBI that have the outcome information for all the samples and were designed for the disease outcome prediction task. In the second task, we developed a workflow (DReCaS – Drug Response Calibration Simulation) that applies gene expression data with gene set enrichment to predict drug response of samples to drugs automatizing the strategy proposed in [14] illustrating its application using liver cancer datasets from ICGC. Both workflows for the two tasks generate a report containing the log of the tasks, the date and time they were executed, the duration of the execution and the memory usage, to monitor the bottlenecks according to the datasets’ size.

### 2.1 ScreenDOP – Outcome disease prediction task

#### 2.1.1 Prediction based on graph kernel matrices

The first approach for the disease outcome prediction relies on the combination of protein-protein interactions with the gene expression data of the patient samples forming the personalized network for each individual [13]. This approach uses numerical features represented by similarity matrices computed from graph kernel functions to perform a screening of the prediction performance according to distinct edge and node attributes. This strategy has three main steps: (i) generation of the personalized network for each patient from the gene expression scores, (ii) extraction of numerical features that represent each patient network, and finally, (iii) disease outcome prediction screening and results exportation.

In the first step, the patient networks is built using the human interactome from the last version released by the HINT database, that contains validated protein interactions [15]. Using these connections as template we filter this network to contain only the interactions whose participants are in the set of DEGs listed in the first column of the masked tables with the log fold change values. For each of the 119526 positive PPIs in HINT we evaluated whether the proteins are both up or down regulated following the cutoff of above 1 (up) and under –1 (down), we use a numeric label for the edge to indicate the up (2) and the down (3) states. The edge is also annotated with the weight parameter composed by the mean of the proteins’ raw expression values. This mean values is set positive in case of up regulation or negative for down regulation. The nodes also have two attributes, the first one is the numeric label that is 1 in case the raw expression value is above 1 and 2 when the raw value is under –1. The second node attribute is the expression containing the normalized raw expression value. We only used the DEG information to prune the interactome and to take a decision on whether or not include a link in order to preserve the the profile observed for each sample and create a variance among the samples’ graph topology.

The numerical features generation from the graphs are computed in the second step through the usage of three combination manners of use node and edge attributes implemented by the kernel functions in Grakel [16]. The performance screening loads the samples’ networks and extracts the nodes and edges attributes in three possible modes only the labels, only the weights or both. The screening encompasses pairwise combination of these three modes for the nodes and edges, forming the dataset passed to compute the similarity matrix of the samples’ graphs representation. From the wide range of choices available concerning kernel functions, we selected three for this screening process, which are the vertex histogram, edge histogram and Weisfeiler Lehman. The criteria used for this choice was the algorithm complexity, avoiding the ones exponential components, and these chosen methods explores information nodes (Vertex histogram) and edges (Edge histogram) separately or both (Weisfeiler Lehman) [17, 18].

The calculated similarity matrix are squared and undergo a grid search parameter optimization of two components of the support vector machine classifier [19], which are the regularization (C) ranging from 10*^−^*^2^ to 10^2^ and the kernel coefficient (gamma) from 10*^−^*^3^ to 10^0^. The report of this optimization calculates and saves the kernel function name and the values of accuracy, precision, recall and f1 metrics [20] for the best model chosen from a 10-fold cross validation using each possible pair of these parameters. The second and last report uses the sample true labels and graph similarity matrices to compute, for each sample, the number of similar (above 0.7) samples that are in the same and in the opposite class to obtain a decision to understand the features distribution prior to the classifier application.

#### 2.1.2 Prediction based on gene set enrichment analysis

The second strategy applies gene set enrichment analysis (GSEA) to retrieve the pathway enrichment scores according to the normalized raw expression values of the samples, using these scores for all pathways found as numerical features for the outcome classification. This strategy is also composed of three steps: (i) numerical features generation; (ii) model training and prediction evaluation, and (iii) Results exportation and post processing analysis.

From the data preparation, the gene names were originally identified according to the Uniprot reference. However most of the gene sets available for enrichment analysis from gene expression data use the conventional HGNC symbol, so firstly we filtered the genes available in the full raw expression values matrix for those that have a corresponding mapping to a gene symbol. In this approach, we use all genes of the dataset instead of filter the DEGs as the first strategy, and the filtered table mapped to hgnc symbol pass by a second filtering step to account for those genes that are indeed present in the gene sets available in the 2021 release from the Kyoto Encyclopedia of Genes and Genomes (KEGG). The tool used for GSEA analysis was GSEApy [21] and the chosen gene sets library was *KEGG 2021 HUMAN*, the gene symbols were collected from each pathway identifier and used for the second filtering process. Since both the raw expression values were already normalized, we submitted them directly to the enrichment analysis and grouped the table by sample, originating a matrix with the pathway scores in the columns and each sample in the rows. We also attach in this matrix the information of sample identifier and the true outcome class in the first columns.

In the second step, the numerical features matrix is used to train the model following a similar hyper parameter tuning using grid search optimization, however in this approach the type of classifier used was the Adaboost that implements an ensemble technique combining decisions of weak classifiers to decide the label for each record. The grid search iterated over two parameters: learning rate from 10*^−^*^4^ to 10^0^ and number of estimators (10, 50, 100, 500) [22]. To deal with the unbalanced number of records for each class, in this strategy we applied the SMOTE (Synthetic Minority Over-sampling Technique) strategy to generate more examples of the underrepresented class [23, 24].

To build the the first results report of the third step of this approach, we also used a 10-fold cross validation taking the mean of the four performance evaluation metrics (accuracy, precision, recall and f1) as well as the standard deviations. Each step of the grid search results is also documented with the mean accuracy and its standard deviation reached by their combinations. The last post processing report generated is a comparison among the gene neighbors from the correlation matrix from the raw expression values and their neighbors from the HINT validated protein interactions. For this report, we used the masked matrix containing only the DEGs and we used a cutoff of above 0.7 and under –0.7 to nominate the respective positively and negatively correlated genes. This report intended to investigate the agreement of the physical complexes with the snapshot of the mRNAs quantified inn the samples. We also stratified this analysis in three cases, among each outcome class separately and considering the whole range of samples.

### 2.2 DReCaS – Drug response calibration simulation workflow

We developed a workflow (DReCaS – Drug Response Calibration Simulation) to automatize the methodology for drug response calibration simulation proposed by [14], adding new functionalities to complement the original method and providing then an end-to-end workflow to perform drug response experiment screening. The workflow comprehends the following procedures: (i) Raw data processing, (ii) Model training, (iii) Weights optimization for the calibrated scoring matrix, (iv) evaluation and prioritization of individual drugs, and (v) drug combination evaluation.

#### 2.2.1 Raw data processing

The data processing procedure treats the raw gene expression counts for each of the samples and starts treating gene nomenclature, to normalize to the standard pattern of HGNC symbol. It identifies whether there are genes identified with Ensembl or Uniprot identifiers and uses the Biomart [25] implementation provided in GSEApy [21] to map the gene identifiers to HGNC symbol. To avoid sending repeated requests for the same genes over future experiments, it builds an internal dictionary that is loaded to enhance the speed of this mapping phase. It also saves a list of the identifiers that could not be found, to exclude in future experiments.

The input expression file is a tab separated file containing three columns representing the gene id, the sample identifier and the raw read count. We transform this table using the pivot technique to generate the standard table with gene identifiers as index, and the raw gene count values for each sample column. The matrix of the mapped genes per samples are then transposed to be applied in the rnanorm tool [26] to generate a normalized final table. The user may choose among five types of normalization methods, which are FPKM (Fragments Per Kilobase of transcript per Million), FPKM UQ (FPKM upper quartile), TMM (Trimmed Mean of M-values), TPM (Transcripts Per Million) and CPM (Counts Per Million). The data transformation for FPKM UQ, TMM and CPM occurs directly, while FPKM and TPM requires the genes length information in order to calculate. The workflow handles the human genome builds 37 and 38, and loads the gene lengths from the Gene Transfer Format (GTF). The mapping is originally available for all genes in ensembl id, but rnanorm expects only the genes contained in the table to be normalized and then this step of the pipeline maps and filters the gene length dictionary.

The normalized table is transposed back to the standard genes by samples format to be applied in the gene set enrichment analysis. The user may choose any of the available gene sets in the library to enrich the data, but there are only 15 gene sets concerning pathway information, most of these are versions of WikPathways and KEGG. This analysis generates the samples pathway scores matrix that will be used for calibration according to the impact of drugs on the identified pathways.

#### 2.2.2 Model training

This procedure is responsible for training the model that differs from healthy and disease samples, and calculates the modified pathway scores from the previous calculated table exactly as specified [14]. However, in the training phase we added a performance log to track all the most used performance metrics values (accuracy, precision, recall and f1), allowing model transparency at every step of the pipeline. The features used in the model are the normalized enriched scores for all the pathways identified, distributed across the healthy and disease sample classes. We use the 10-fold cross validation strategy implemented internally in the Elastic Net regressor [27, 28], selecting and saving the model with the highest precision mean. In case one of the classes is poorly represented (under 40%) by the available samples, the workflow automatically applies the SMOTE oversampling technique to treat the imbalance [23, 24].

#### 2.2.3 Weights optimization for the calibrated scoring matrix

This procedure uses an optimization strategy to get the best values of the three weights for a maximum of 100 rounds of evaluation using the optuna optimization framework [29]. The optimization experiment is configured to setup the three weight values ranging from 1 to 30 as integer values., using the trial values of the round it computes the modified version of the sample pathway scores, following a user specified gold-standard drug list. This list may be composed of approved drugs (using Drugbank identifiers) or drugs that are being evaluated in clinical trials for the disease inherent to the dataset. The evaluation function builds the score of each trial by firstly calculating, for all gold drugs, the number of original disease samples that were classified in the opposite class (healthy) by the trained model in relation to all disease samples. This ratio is then used to compute the number of drugs in the list that changed at least 70% of the disease sample labels, forming the final score based on the ratio between this number and the total number of drugs. The goal is maximizing this score with the weight values, since these drugs were approved or passed to the final phases for the disease treatment. The final values of the best solution are saved to be loaded in the previous procedure or the next procedures of drug or drug combination ranking and prioritization.

#### 2.2.4 Evaluation and prioritization of individual drugs

The first step of this procedure is calibrating the samples pathway scores according to the drug influence in these pathways through their target genes. Although the mathematical method remains the same, we used algorithm strategies of data indexation to efficiently compute with a fair memory and cpu usage, for a considerable number of drugs. Before the calibration, two pieces of information are calculated, which are the mean score of the influence of the drug in the pathways present in the samples pathway score matrix. We use the table with the positive or negative correlation^3^ of a drug on modulating a specific gene, and get the mean from the genes in the intersection of the two sets. The second information is generated summarizing the pathway scores by sample class groups using the average and taking the absolute value of the mean difference among healthy and disease samples.

One of the new features is the possibility to use models trained from another experiment, skipping the training step, that is activated only when the mapping file connecting the sample identifier and the respective class is available. The transfer learning mode, the absolute mean differences are inherited from a previous experiment to perform the calibration. The calibrated matrix does for a given pathway of a sample only occurs if the drug score for that pathway is distinct from zero, otherwise its signal (positive or negative) is used to modulate the weights assigned to the sample original score. There are three possible weights that depend on the position of the current pathway absolute mean difference (*pdf mean*) in relation to the quartile values from all the absolute mean difference list. In case the user provides a file specifying the values of each weight, it is applied in this step, otherwise, it uses 20 (*pdf mean* above third quartile), 5 (*pdf mean* above second and under third quartile) and 10 (*pdf mean* under second quartile). By default, as this step occurs prior the optimization and the trained model is required for weights optimization, it computes for all drugs available in the drug-gene relation file using the default weight values.

Then, all the drugs that have a positive or negative modulation in a certain gene that belongs to one or more enriched pathways for the provided dataset are evaluated according to the trained model. From the second procedure, the calibrated pathway scores for a specific and single drug were already ready to be applied as features in the model. We use a similar evaluation criteria mentioned in the weights optimization, but it computes only the ratio of disease samples whose modified scores lead to change the model decision for each drug being evaluated. This score is then applied to generate the ranking report ordering the drugs from the highest to the lowest ratio. The second report is a binary matrix indicating whether a specific sample obtained a favorable response to a certain drug in the list.

In the transfer learning mode, the workflow requires the samples pathway score of the original experiment of the model used, so that it can calculate the number of features expected by the model. The original experiment of the model and the target experiment must use the same gene set to calculate the features correctly. In case some pathways of the target experiment are not found, the features array is cut to fit the number required by the model, otherwise, it completes with zeros. To leverage the maximum information, a mapping is made from the target enriched pathways and the model original experiment ones, those that are not found receives a respective pathway that contains the highest intersection of genes in common.

#### 2.2.5 Drug combination evaluation

The last procedure allows the user performing a similar screening as explained in the previous procedure, using the same evaluation criteria, but with a customized list of drug combinations. To accelerate the calculus of the calibrated sample pathway scores, we redesigned the algorithm strategy, splitting the signal identification from drug scores by pathway and the identification of which weight to use based on the absolute mean differences to atomic functions so that the computation of the modification is mapped for all drugs used for each combination and then later aggregated into the original sample score. This computation method forms each sample numerical features that are evaluated by the model. It generates the same above mentioned reports but accounting for the grouped drugs instead of single drug identifiers.

### 2.3 Datasets for evaluation

#### 2.3.1 Disease outcome prediction task

We used two publicly available gene expression datasets in the experiments corresponding to the Leukemia (accession GSE425^4^) and Ovarian (accession GSE140082^5^) cancer types. These datasets were extracted from the GEO DataSets (GDS) database [30] from the National Center for Biotechnology Information (NCBI), while the leukemia cancer dataset used the micro array sequencing technology, the ovarian cancer one dataset was generated through RNA-Seq high-throughput technology. Both dataset contains labels informing the sample outcomes (dead or alive) and were previously applied in survival analysis [31, 32].

The ovarian cancer dataset contains 380 samples, which are subdivided into four groups according to the cell sub type (mesenchymal (39 treated / 34 control), proliferative (47 treated / 50 control), immunoreactive (70 treated / 54 control) and differentiated (43 control and treated)), and in each group a portion ranging from 48.45% to 56.45% of the samples were treated with the Bevacizumab drug while the other half corresponded to the control samples. The raw read counts table were available for all the samples and the genes were provided using the illumina platform identifiers, that were mapped accordingly to the protein identifiers in the Uniprot database [33]. We performed the differential gene expression analysis using the DeSeq2 tool [34] in these four groups and extracted the log 2 fold change measure to apply to the patient sample networks. The experimental design matrix was derived from the metadata provided in the information page of the GEO dataset as well as the raw counts for each sample and genes. The raw gene expression counts were normalized using the FPKM (Fragments Per Kilobase Million) upper quartile technique as it is the standard normalization provided in databases such as TCGA [35].

The Leukemia dataset contains 119 samples from 65 peripheral-blood samples and 54 bone marrow specimens from 119 adult patients with Acute Myeloid Leukemia provided by the AML Study Group Ulm (Ulm, Germany) [31]. The patients underwent to two treatment protocols (AMLSG-HD98A and AMLSG-HD98B) described in [36] and the original study performed a gene expression profiling to find out though the screening of supervised and unsupervised learning strategies predictive genes that account for survival. They established a correlation among the gene expression and survival outcome. The gene identifiers were mapped according the GEO platforms tables related to the samples that provides the HUGO Gene Nomenclature Committee (HGNC) symbol for each gene id of the individual sample expression information. The raw gene expression values used to build the patient networks for each gene in this case was the log2 ratio found in the last column of the sample matrices (provided according to their GSM (GEO Sample) accessions) containing the quantitative data for the micro array, indicating the log2 ratio of the mean among the channels. As this dataset was derived from microarray technique, we obtained the normalized fold change of the differential gene expression using the limma tool [37].

From these two strategies, three standard files are derived to be used in the in the methods proposed along this article, which are the matrix containing the raw normalized expression of the genes for each sample, a masked matrix containing only the differentially expressed genes identified with their values of fold change filtered according to p-value under 0.05, the last file is a table with two columns informing the sample identifiers and their true outcome. All the gene identifiers from these datasets were transformed to Uniprot identifiers.

#### 2.3.2 Drug response simulation task

The original version of the method for drug response simulation [14] tested the strategy on four TCGA datasets that comprehends four types of cancer, which are Breast Invasive Carcinoma (TCGA-BRCA), Prostate Adenocarcinoma (TCGA-PRAD), Liver Hepatocellular Carcinoma (TCGA-LIHC), and Kidney Renal Clear Cell Carcinoma (TCGA-KIRC). In this paper, we focused the experiments on two ICGC datasets concerning liver cancer that are LIRI-JP^6^ and LIHC-US^7^. Besides the type of data analysis is the same as the first task, the experimental design of the datasets used in the first task are not divided in normal and disease samples, this fact would lead to a misinterpretation about the predictions, since some tissues of those that survived could also be extracted from disease samples. The LIRI-JP project contains 243 disease samples and 202 healthy liver tissue samples. Although we focused specifically in the gene expression dataset, this dataset publicly provides other omic types of datasets such as simple and structural somatic mutations. copy number variations. We used the samples metadata to build the labels file to build the model in the drecas workflow. We only used one type of cancer to illustrate and discuss in details the behavior of the model and drug ranking according to the pathway gene sets and types of normalization. The LIHC-US project contains 297 healthy and only 48 disease samples present in the gene expression file, so this dataset has a huge imbalance. This dataset was then applied as the target to test the transfer learning function of our workflow and to test the performance applying the oversampling strategy. In this case, simple somatic mutations, copy number variation, methylation, micro RNAs datasets were also available.

## 3 Results

### 3.1 Disease outcome prediction

#### 3.1.1 DEGs analysis in the datasets

According to the limma tool results, there was 6686 differentially expressed genes with a significance under 0.05 in the leukemia dataset. While in the ovarian dataset, combining all the DEGs resulting form the DESeq2 tool for the four cell subtypes experiments, the total number of DEGs was 1842. In each dataset, a considerable number of protein interactions were reduced from the original HINT network, from the total number (119526) of positive PPIs from HINT, the leukemia base interactome filtered with the the differentially expressed genes contained only 28573 (23.9%) protein pairs. The loss observed in the ovarian cancer dataset was larger than the one in leukemia dataset, maintaining only 1382 protein interactions (1.16%).

#### 3.1.2 Sample networks characteristics

To evaluate the first strategy based on the features generated from the sample protein networks, we calculated the distribution of node and edge labels across each dataset since the kernel functions applied take this information into account. In both scenarios, the summary of the network analysis of the samples showed that the 99% of the complexes or protein pairs were formed by proteins that were up regulated at the same time and this proportion is equal in the two classes of samples. However in the leukemia dataset 34% of the node labels were down regulated and the rest was up regulated. In the ovarian cancer dataset, using the same cutoff parameter, all the nodes were up regulated as its edges.

The number of nodes (5590) and edges (28485) were the same for all the samples since that were only one outcome of DEGs. In the ovarian cancer scenario, there were four design matrices according to each subcell type, that generated distinct number of nodes and edges for the samples belonging to each subcell type (Table 1), but in general the variation for nodes (9) and edges (64) were small (64 for edges and 9 for nodes), specially for the number of nodes.

**Table 1.** Number of nodes and edges derived from each experimental design matrix in the ovarian cancer dataset.

| Subcell Type | Number of Nodes | Number of edges |
| --- | --- | --- |
| Immunoreactive | 807 | 1369 |
| Proliferative | 806 | 1365 |
| Differentiated | 798 | 1305 |
| Mesenchymal | 803 | 1350 |

The other attribute aggregated into the sample networks is the weight and can also be used alone or combined with the label in the kernel functions. The distribution of node and edge weights showed in figure 1 indicates that the raw expression values was not enough to create a variance among the samples belonging to distinct classes, apart from the outliers, the major portion of the data points are concentrated equally along the quartiles for the ovarian cancer. However, in the leukemia dataset, the outliers were also observed but in the dead class samples instead of the both classes as observed in the ovarian cancer ones. Although the majority of data points were slightly changed around the mean values comparing the two classes, there was an improvement in the weight variance in relation to the ovarian cancer dataset. Interestingly, limma and DeSeq2 showed that both datasets have the majority of the pairs with positive weights for the edges. The dataset derived from microarray shows that the raw normalized expression values (the weights assigned to the nodes) are mostly negative.

**Fig. 1.**
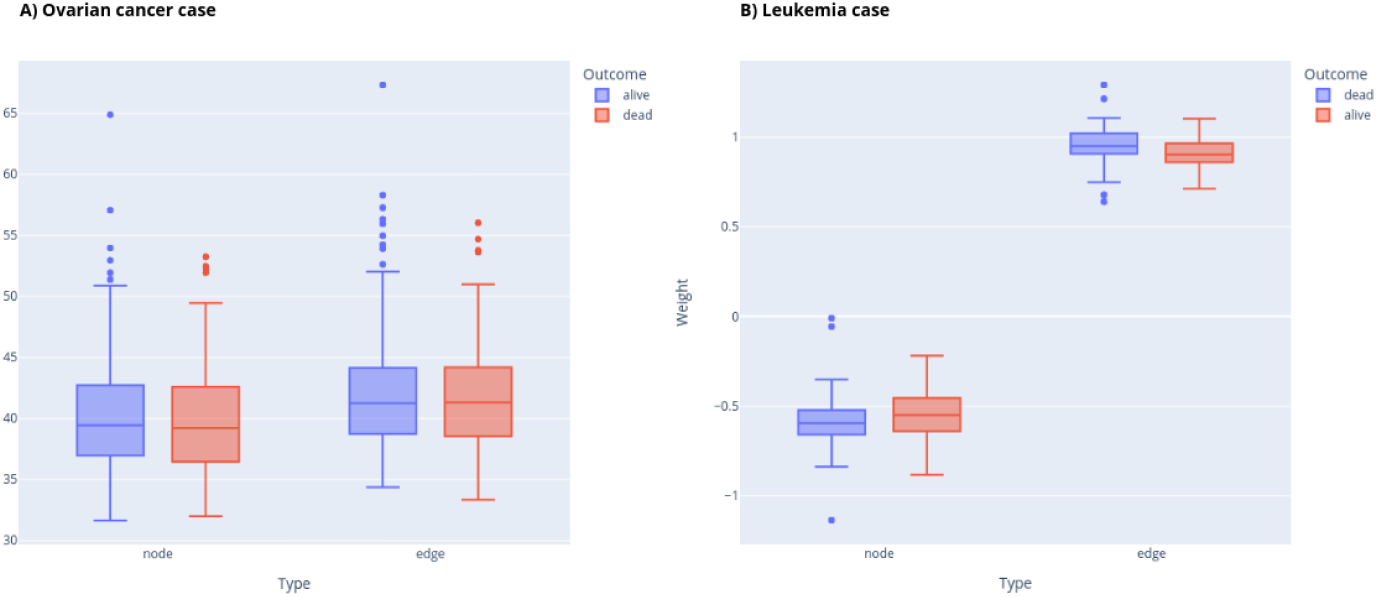
Comparison of the node and edge weights distribution in each scenario. Scenario A represents the Ovarian cancer data (blue bar is the alive class and the red one identifies dead class) and B presents the leukemia data (in this case, the order changed, so blue bar is the dead class and the red one means the alive samples).

Other studies have proposed the integration of gene expression data to enrich networks with protein interactions from the DEGs found [38] to understand the modulation in disease scenarios. Here, we created a framework that allows the systematic simulation of sample-specific protein interaction architecture to understand the modulation using multiple combinations of node and edge attributes. These networks may be used to combine the treatment/control information on pruning the networks and allows represent the sample specific expression levels on the nodes, amplifying the representation of sample information. The existing approaches [38–40] creates a general protein interactions network and the proceed to enrich with other functional databases such as KEGG or Gene ontology, without taking into account the individual sample networks.

#### 3.1.3 Performance evaluation of the network-based strategy

As expected from the variations found among the attributes in the Leukemia and Ovarian cancer datasets, the performance evaluation metrics of the models achieved up to 80% of precision in the Leukemia. The best parameters found for the classifier grid search in this dataset were gamma equivalent to 1, and the C parameter depended on the kernel function, being 0.01 for *EdgeHistogram* and 10 for *WeisfeilerLehman* [18]. These two kernel functions obtained the best results for all main metrics (f1, precision, recall and accuracy), but the highest values remained in the precision and f1. Still in the leukemia dataset, the top ranked results of the screening showed that the attribute weight for the node was predominant and, among these, the combination with the edge label was mostly found.

However, in the ovarian cancer dataset this strategy was not successful, as the network analysis showed that the attributes have almost the same distribution in both classes, this was reflected in the performance metrics, the model was not able to learn how to differentiate the samples. The values chosen for the C and gamma parameters were 0.01 and 1 for all combinations in the screening, although there were occurrences of accuracy above 65%, the other three metrics did not achieve values above 34%. The best values reached followed the same pattern found in leukemia, with the weight attribute for node and edge combined with the *EdgeHistogram* and *WeisfeilerLehman* kernel functions.

A previous work also proposed the combination of gene expression and protein interaction into a network and the usage for disease outcome prediction for cancer disease using kernel matrices [13]. We not only allowed transparency on the features engineering but allowed flexibility on the samples network architecture, but we also handled the data processing extending their application to gene expression data from RNA-Seq technology.

We also investigated the hypothesis of bypassing the grid search and classifier application and derive the most the sample neighbors using only calculated similarity values from the kernel matrix. Using the specified cutoff of 0.7, as expected the experiments using only label as node attribute values, could not discriminate the samples from a different or same class, it marked all the samples as similar. Using the weights, the separation among those from the opposite class were more accurate, agreeing with the best models for the leukemia scenario. In the ovarian cancer scenario, 66% of the experiments generated kernel matrices whose pairwise similarity were greater than 0.8, while the weight contributed to enhance the prediction in leukemia scenario, in the ovarian cancer all the experiments (33%) produced similarity values under 0.6 even for those graphs from samples of the same class. These results demonstrate that the similarity matrix alone is not a good source to rank the sample graphs. A similar analysis was performed by [41] that used the comparison among the patient graphs using kernel function based similarity matrices, and then proposed ranking these patients according to the phenotype under investigation. Their goal was classifying those that will follow to a deep evaluation to assess the correct prognosis, they do not rely completely in the kernel matrix to check the patients that group together with those from the same phenotype.

The experiments execution log showed that this strategy took a total of 5 seconds to run all the steps for the leukemia dataset and 15 seconds for the ovarian cancer that has about three times leukemia samples number. Apart from the first step, all the other were executed in less than 0.7s for leukemia and 1.4s for ovarian cancer. The most consuming task was the generation of the personalized sample networks, that took 5.46s for the ovarian cancer and 3.6s for leukemia dataset. This task is also the one that required at most 55.33Mb and 41.25Mb, for ovarian cancer and leukemia datasets respectively.

A recent review describes in details the role of the variety of graph-based machine learning methods for disease prediction [42], including the ones based on kernel functions and support vector machine that was used in our strategy. They pointed as a future direction strategies to create an effective graph as a potential research direction, by allowing the automatic simulation of samples networks with distinct weights and attributes we move a step towards attending this research direction. We allow these simulations in computationally efficient manner while performing a screening easily configured by the user.

#### 3.1.4 Performance evaluation of the GSEA pathway scores strategy

We first evaluated the fifteen most enriched pathways identified for each Scenario, in order to access the score distribution of the pathways across the distinct classes (Figure 2). The Leukemia case obtained 278 enriched pathways, and the top 15 pathways were coherent to the since most of these are related to diseases in involving immune system aspects, such as Diabetes mellitus, autoimmune diseases and immunodeficiency. The values of these top pathways were similar among the two classes and for Autoimmune Thyroid disease the mean values were the same. In the ovarian cancer scenario, some general pathways like ribosome, spliceosome and RNA degradation among the most enriched ones. There were two interesting cases in this scenario, the nicotine addiction pathway was negatively enriched in the alive group while in the dead group it was almost zero. The maturity onset diabetes pathway was negatively enriched for both groups of samples, still the dead group value (–350) was higher than the alive one (about –500). While the ribosome pathway were positively enriched in the alive group, it was negatively represented in the dead group.

**Fig. 2.**
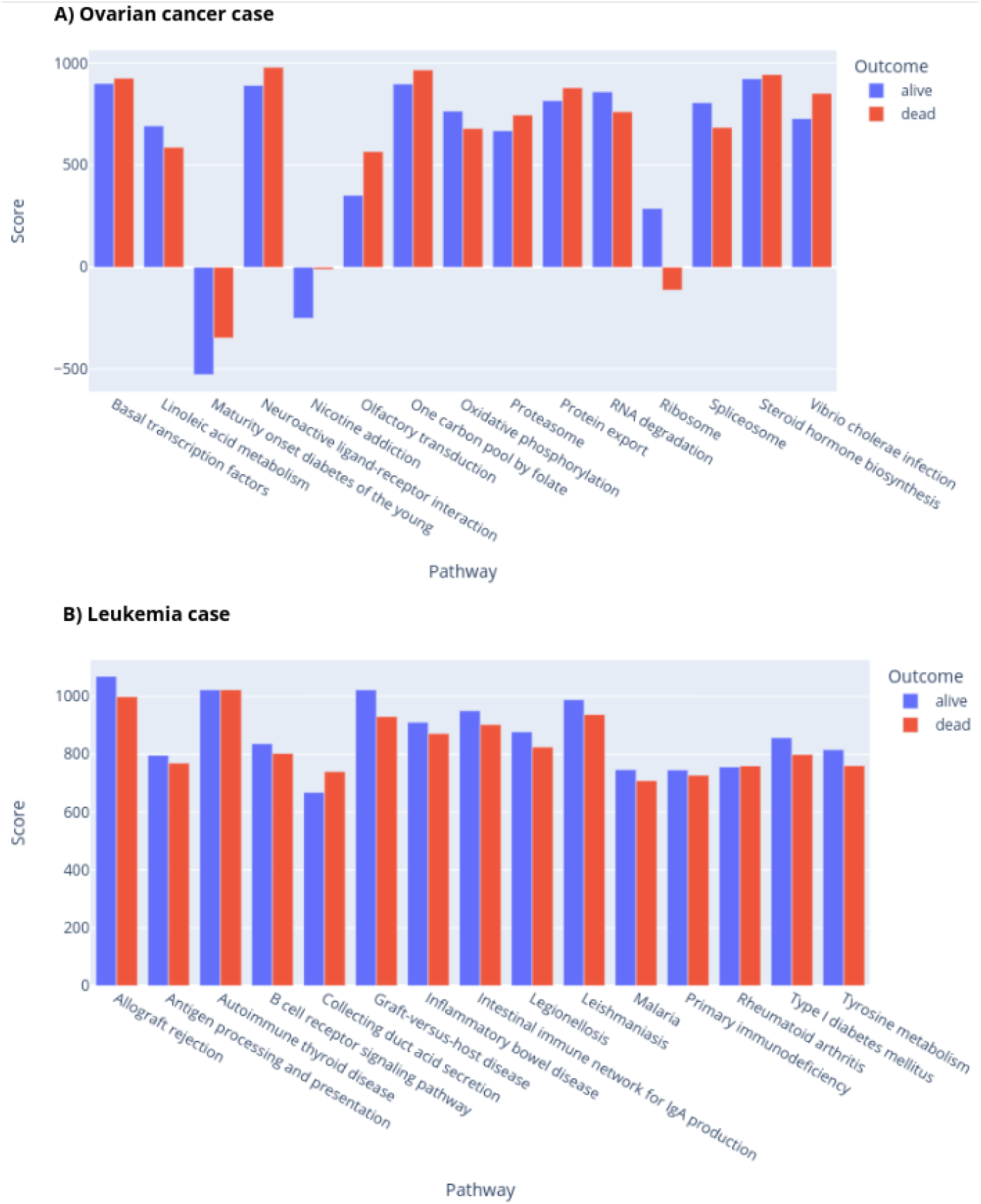
Comparison of the fifteen most enriched pathways of each disease scenario, grouping the values obtained by the samples according to their class (dead or alive) according to the average score.

We evaluated the prediction comparing the performance evaluation metrics using the oversampling balancing method before passing to the 10-fold dataset split, and passing only the original samples. According to the grid search parameter tuning for Adaboost, the best accuracy (reaching 80%) were yielded by learning rate values set in 1 and 500 estimators for the ovarian cancer scenario and all the top ranked parameters came from the balancing experiments. The other balancing strategy yielded a better result (64.7%) for the leukemia scenario, in this case the favorable learning rate was 10*^−^*^3^ combined with 50 or 100 estimators. Regarding the performance evaluation metrics (Figure 3), while the model was clearly improved by the usage of the SMOTE technique, not only for the accuracy but mainly for the equilibrium among the other three metrics (precision, recall and f1), in the leukemia dataset, the balancing decreased the values of all these metrics, being the greatest reduction observed in recall (–0.36) and f1 (–0.21).

**Fig. 3.**
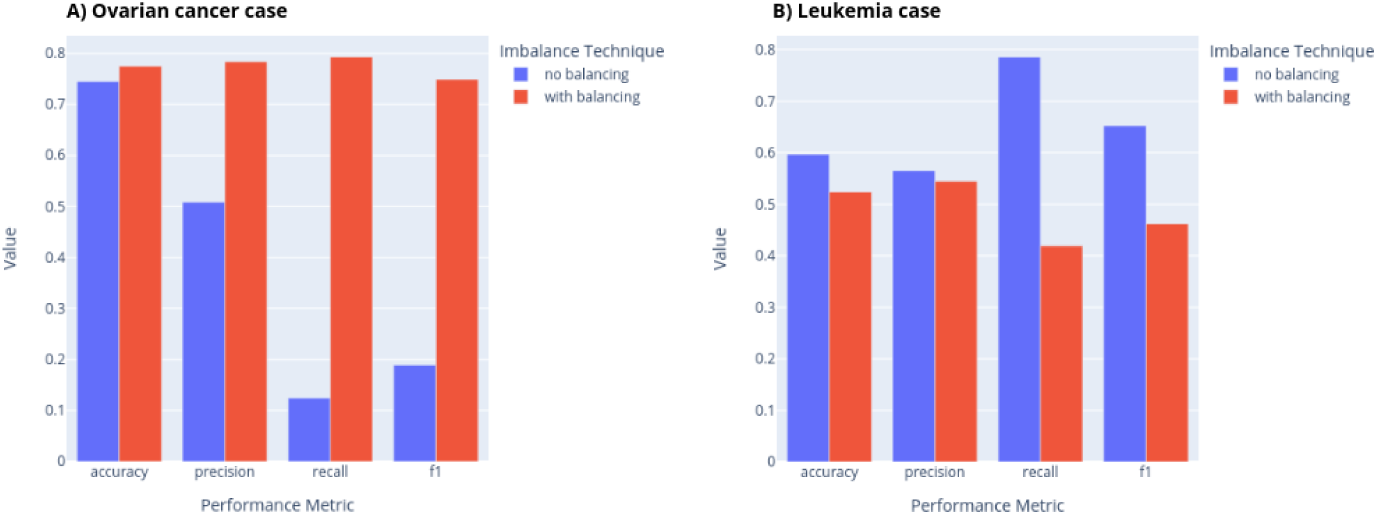
Comparison of the performance evaluation metric values in the leukemia and ovarian cancer scenarios using or not the SMOTE balancing technique in the samples’ data.

In this strategy, we introduced the representation of the gene expression profile using pathway scores from gene set enrichment analysis to test the ability to predict disease outcome as a domain-agnostic predictive method. Other published works proposed methods that are highly domain-dependent and require human intervention for curation [43], this method also uses GSEA but only as a post processing analysis tool. Other methods also used network-based approaches for outcome prediction with gene set enrichment [44, 45] but their focus was using the gene set enrichment as a post processing strategy to validate the findings across other biological data sources. Our purpose with the strategy presented is using the pathway scores from gene expression to directly predict the prognosis, while the gene network built by these methods aims to extract individual gene signatures that could potentially improve prognosis prediction.

Concerning the computational efficiency (Table 2), this strategy steps take more time to execute in relation to the first strategy presented in the previous section. In the leukemia scenario, the step that trains the model and performs the parameter tuning took the longest time (99s), while the features generation (step 1) took 15s, it used the highest amount of memory (6.8Mb). In the ovarian cancer scenario, the first and fastest step took 40.4s and promoted a memory usage of 166Mb, and the most time-consuming was the report generation (step 3) spending 770s (almost 13 minutes).

**Table 2.** Comparison of memory usage and execution time among the datasets for each step of the GSEA-based strategy.

| Step | Leukemia |  | Ovarian cancer |  |
| --- | --- | --- | --- | --- |
|  | Time (s) | Memory (Mb) | Time (s) | Memory (Mb) |
| 1 - Features generation | 15 | 6.8 | 40.4 | 166.9 |
| 2 - Model training and evaluation | 99.63 | 0.073 | 240.9 | 2.4 |
| 3 - Report of PPI vs Correlation | 42.78 | 1.18 | 770.78 | 7.62 |

Following the report generated by the third step, in both scenarios, the network based in negatively or positively correlated proteins from the raw normalized expression values almost no overlapping with the physical complexes of the protein interaction network. Indeed, it is known that proteins that interact to each other tend to present stronger correlation in in the gene expression analysis [46], however there were few neighbors of the proteins from PPI in the correlation matrix from gene expression, explaining the poor overlapping observed. Only around 7% of the proteins obtained at least one neighbor which was also present in the protein interaction network in both disease scenarios. Table 3 shows that in both scenarios, the highest concentration of correlated genes occurred in the the sample set of the dead outcome. Only in this outcome there were negatively correlated genes for the ovarian cancer scenario. This report was designed to understand the relationship among the enriched pathways and the gene expression correlation across phenotype-specific sample groups, and extract possible interesting deviations. As we showed in the table the bad outcome yields expressive increase of genes correlated to each other showing potential dependencies from each other to guide personalized medicine [47].

**Table 3.** Comparison of the number of positively and negatively correlated genes considering three sample sets: all, only dead, only alive.

| Sample set | Leukemia |  | Ovarian cancer |  |
| --- | --- | --- | --- | --- |
|  | Corr. (+) | Corr. (-) | Corr. (+) | Corr. (-) |
| All | 1211 | 18 | 4530 | 0 |
| Dead | 2986 | 398 | 7414 | 46 |
| Alive | 1885 | 44 | 4856 | 0 |

### 3.2 Drug response calibration simulation

#### 3.2.1 Assessment of normalization types and pathway gene sets effects

Since the core method for drug response simulation was already extensively tested in [14], we evaluated the new functionalities proposed in this paper. Firstly, we made two experiments in the LIRI-JP dataset to test (1) all the types of normalization and (2) all the recent versions pathway gene sets. The evaluation criteria was based on two aspects: the roc auc and precision metrics to evaluate the model training; the distribution of the changed label ration when testing with current drugs that are on clinical trials for liver cancer. While changing the gene set, we fixed the normalization type as the one used in TCGA datasets (FPKM UQ) with the default weights proposed in [14] (20, 5 and 10). The same protocol was adopted while changing the normalization type, using the KEGG 221 Human as default.

The results for normalization type (Figure 4) show that all the tested methods had a mean precision close to one, however their performance following roc auc metric varied around 0.75, with a great variance around the mean for cpm, tmm and tpm. The least variance occurred using fpkm uq, with a mean value of 0.74. The methods with the minor values of mean for roc auc were the same that also have the lowest values of changed label ratio. Besides all the methods yielded rations above 0.8, fpkm uq and tmm obtained similar mean values around 0.93. Indeed, fpkm uq provides the most robust result.

**Fig. 4.**
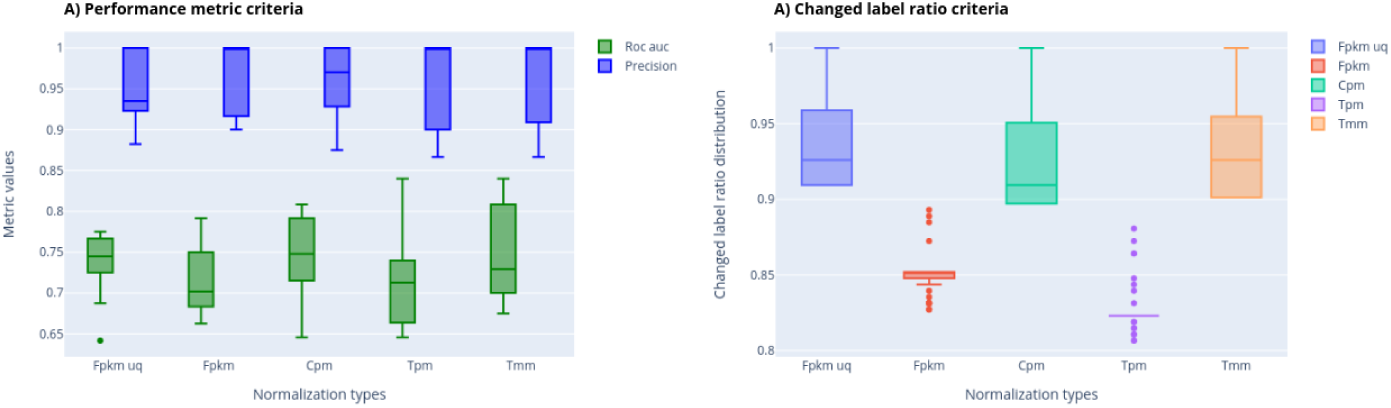
Evaluation of all types of normalization (fpkm, fpkm uq, cpm, tpm, tmm) according to the roc auc and precision metrics for the model quality criteria, and following the changed label ratio on calibrating by clinical trial drugs.

The gene sets evaluation results (Figure 5) demonstrated that except for the pfocr, all the gene sets obtained a mean value of roc auc above 70%. Wikipathways obtained a great variance around this mean value besides being similar to kegg (around 0.75). Combining the results of changed label ratio, only kegg and wikipathways obtained a mean value above 80%, while the elsevier and pfocr obtained an unsatisfactory performance above 0.6.

**Fig. 5.**
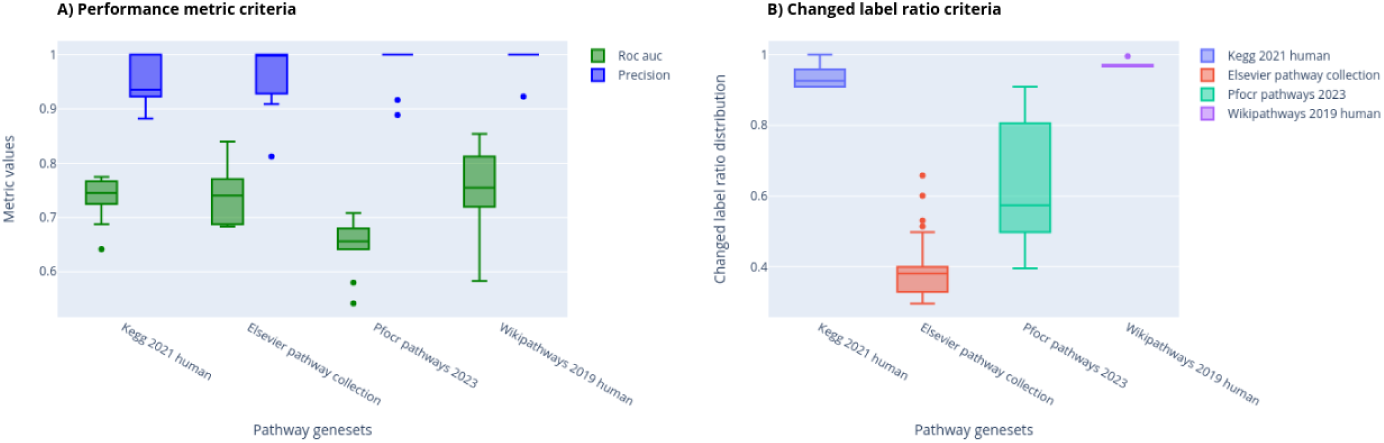
Evaluation of all types of pathway gene sets (pfocr, kegg, elsevier, wikipathways) according to the roc auc and precision metrics for the model quality criteria, and following the changed label ratio on calibrating by clinical trial drugs.

#### 3.2.2 Transfer learning evaluation

We trained the model in LIRI-JP and applied this model to evaluate on the LIHCUS samples and processed both datasets using the fpkm uq normalization type and the kegg geneset, taking advantage of the screening results presented in the previous section.

We extracted a list of 150 drugs that went on clinical trials for liver cancer neoplasm (mesh identifier 68008113) ^8^, uniting this list with approved drugs^9^ for this type of cancer. This list was used to calibrate and optimize the scoring matrix weights, after 100 trials, the best highest percentage of these drugs that changed 80% of the disease sample labels were around 12%, found in the second trial, resulting in the final weights of 1, 27 and 20.

An important point for transfer learning application is the compatibility of the enriched pathways. Although we projected the workflow to treat the possible gaps among the features in the target data and those that form the original model, we checked the enriched pathways in common among these two datasets and 304 out of 305 pathways in LIHC-US were contained among the 305 pathways in LIRI-JP. The *Pathways in cancer* pathway was only found in LIRI-JP.

Firstly, we evaluated the transfer learning mode using the models of the two experiments (LIRI-JP and LIHC-US) to predict the class of their own samples and each other ones. Figure 6 shows that even though the LIHC-US model was built partly using synthetic data to handle healthy class under representation, its performance was very similar mainly for accuracy in relation to the complete model trained on LIRIJP. The roc auc and precision, however, illustrates that indeed LIRI-JP obtained a satisfactory behavior when transferred to other unseen data from other experiment, reaching a precision of around 98%. Both models obtained metric values closer above 90% on predicting their own sample classes, but LIRI-JP based model generalized the learning better than the LIHC-US.

**Fig. 6.**
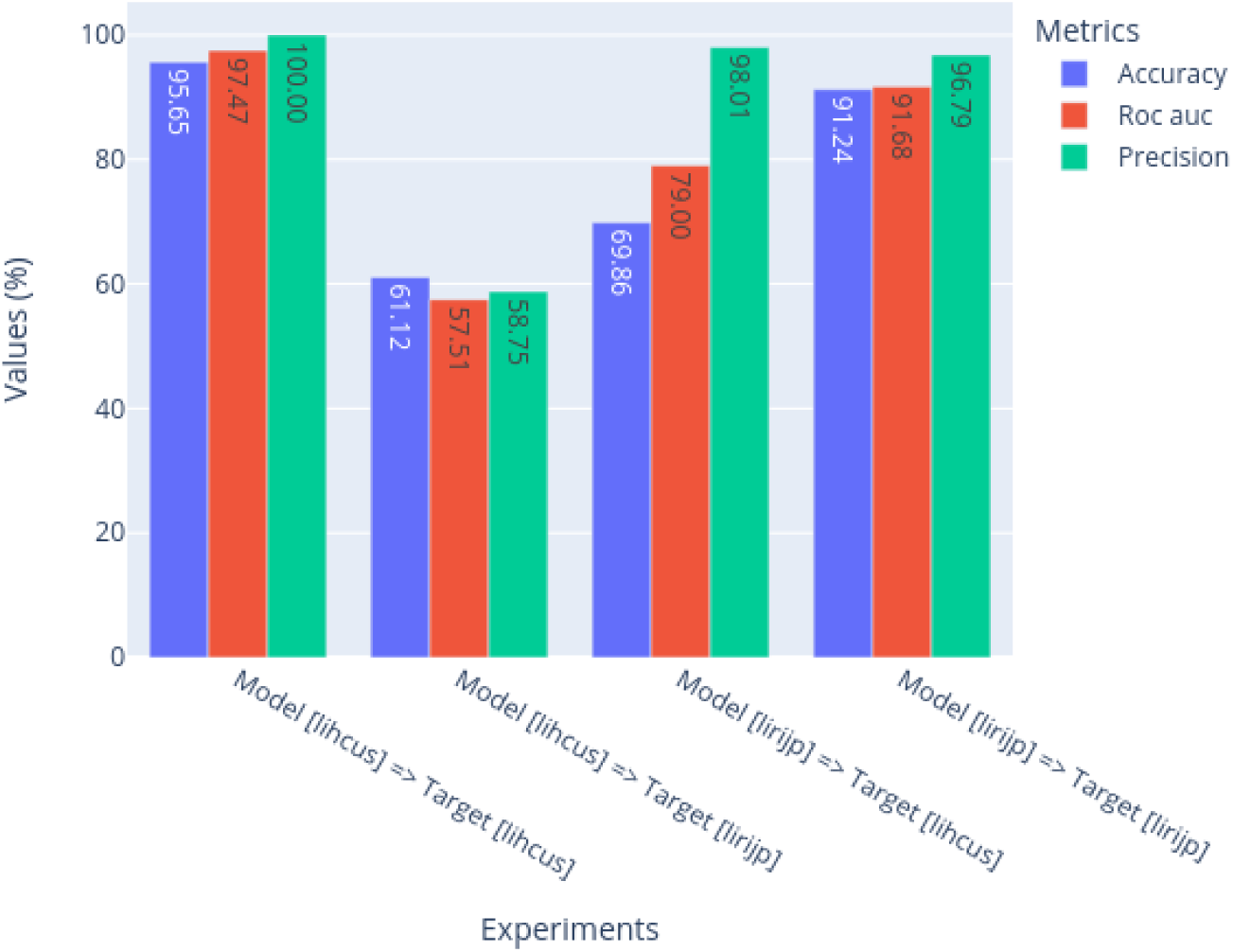
Evaluation of the model performance in a pairwise combination of original models and the targets, considering LIRI-JP and LIHC-US experiments.

As the workflow produces a report about the computational efficiency concerning the execution, table 4 demonstrates that the most time-consuming tasks are the weights optimization and the full screening of 1350 drugs in an individual mode, these tasks reached the maximum values of ~55 minutes (LIRI-JP) and ~49 minutes (LIHCUS in transfer mode). In relation to the memory usage, the only step that requires more than 1GB of ram memory space is the data processing because the raw expression values data loading, in the first step of the data processing it is transformed and discharged from memory, only occupying a large size for a few seconds.

**Table 4.**
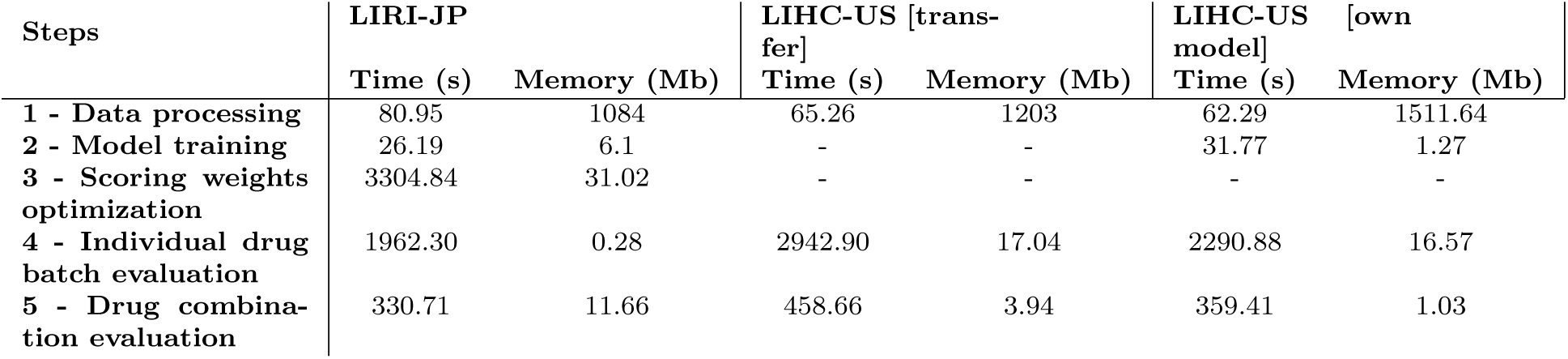
Execution time and memory usage for each of the five steps contained in DReCaS workflow. We tested the execution for LIHC-US in two modes, which were generating its model by oversampling the healthy samples and using the model generated by the LIRI-JP experiment. We skipped the steps 2 and 3 for the second LIHC-US experiment and only the weights optimization step for the first.

In relation to the individual drugs evaluation, we continued comparing the three experiments (LIRI-JP, LIHC-US transfer and LIHC-US model) cutting the derived ranked list using a threshold of 80% of the changed label ratio. The agreement results among the drugs in these filtered lists across the experiments pairwise combination showed that the model trained on LIRI-JP found 221 top ranked drugs out the origjnal 1350, while the model trained in the adapted set in LIHC-US only found 71. Using the LIRI-JP model in the lIHC-US samples, the model increased the original number to 130. Even though, more than 90% of the top ranked drugs for both LIHC-US experiments were the same found in LIRI-JP list.

A detailed analysis exploring the ATC (Anatomical Therapeutic Chemical) classes (Supplementary figure 1) represented by the drugs in these lists showed the presence of two drugs classified as antineoplastic agents which are the Arsenic trioxide^10^ (drugbank id: DB01169) and Gilteritinib^11^ (drugbank id: DB12141), the last one is also classified as protein kinase inhibitor. Both drugs are indicated and used to treat Accute Myeloid Leukemia, however recent studies have shown that the Arsenic trioxide induces apoptosis and inhibits the growth of human liver cancer cells [48]. [49] corroborates these findings by demonstrating that inhibits liver cancer stem cells differentiation and metastasis targeting the SRF/MCM7 complex, while the work [50] confirmed this result through inhibition of LIF/JAK1/STAT3 and NF-kB signaling pathways. Gilterinib correlation with liver cancer is not straightforward, according to [51] this medication is used specifically for FLT3-mutated by FLT3 inhibition in Accute Myeloid leukemia, but this work also demonstrates evidence of an off label usage of the liver cancer’s approved drug sorafenib. The mono therapy with this drug in a group of patients induced durable remissions presenting allo-immune effects.

We tested the drug combination evaluation using the same methods and the changed label ratio metric, among the true positive pairs of drugs, we used 1 combination cited in [14], eight proposed by [52], and the combination formed by Arsenic trioxide and sorafenib. This last combination is found to promote the apoptosis by upregulating a TNF-related ligand [53]. We randomly generated one hundred distinct drug pairs and all the drug combinations in the true positive set obtained a ratio above 80%.

A previous published method also took advantage of specific cancer cell lines curated information and applied the model trained on this data to evaluate unseen new samples to test transfer learning on drug response in the test set [54]. Although they evaluated using a wide range of parameters and classifiers, our approach is disease-agnostic, since it relies only on the raw gene expression values, and also returns reports of the drug intervention for the pathways of each sample. Furthermore our approach also allows the screening of drug combinations.

## 4 Discussion

We have presented two workflows (ScreenDOP and DReCaS) that handles efficiently and uses only transcriptomic data based on gene expression profile to tackle two biomedical research tasks, which are disease outcome prediction (DOP) and drug response simulation. The ScreenDOP provides two strategies for DOP, and both strategies can be customized allowing the users to perform distinct experiment setups with their own gene expression data. Their parameters for node and edge attributes in the first strategy as well as the balancing option of the second one was tested in two scenarios, using micro array and RNASeq data sources. The DReCaS workflow extended the methodology proposed in [14] by allowing the scoring matrix optimization and transfer learning from one experiment to another. We also standardized the procedure in a reproducible manner dealing since the raw expression values processing till the individual drug screening or custom drug combination response simulation. Both workflows are publicly available with a documentation of the configuration file needed and running examples. They are easily configurable and all the analysis datasets are organized in order to contribute with the data flow transparency and traceability.

In the disease outcome prediction, We showed the relationship among the distribution of features derived from both strategies (samples’ weighted ppi network and enriched pathway scores) and the later prediction evaluation results. The leukemia dataset derived from microarray technique was favored by the first strategy while the ovarian cancer dataset (derived from RNA-Seq) obtained its best performance results using the GSEA-based strategy. The usage SMOTE technique to treat imbalance leveraged the model in the ovarian cancer scenario for all the evaluation metrics, while in the other scenario it was observed a small reduction of these metrics. In both strategies, the accuracy values for the Leukemia scenario surpassed the prediction obtained in [13]. We also showed that our two proposed strategies are fast and robust, spending at most 15 minutes to handle all the procedure. We added two proof-of-concepts, demonstrating that it is not enough using only the graph similarity matrix to check the closest samples and then derive a classification. And we also showed that the neighbors of the gene correlation network have few but most of the times no overlapping with their neighbors in the protein interaction network, in these two scenarios.

Concerning the second task of drug response simulation, we took advantage of the custom configuration to handle multiple experiments in the DReCaS workflow architecture, and performed an experiment screening to demonstrate the behavior of the approved drug indication following distinct normalization types and pathway gene sets. We chose the best combination of normalization (FPKM UQ) and gene set (KEGG), and presented the results of the weights optimization and the influence of the model built in a complete liver cancer dataset (LIRI-JP) in a dataset containing mostly disease samples (LIHC-US), illustrating the transfer learning functionality. The best model could successfully predict the disease samples of the LIHC-US dataset with an accuracy close to 70%. The features could be adapted and the drug prioritization for datasets concerning the same disease were in agreement. We found a promising drug originally used to treat leukemia as a top ranked drug from our screening results and validated as a potential liver cancer therapy [48–50]. Our workflow can be applied to test multiple drug combinations enhancing drug repurposing and leveraging personalized medicine application.

## Code availability

The ScreenDOP code is available in a Github public repository at https://github.com/yascoma/screendop while the DReCaS is available at https://github.com/YasCoMa/caliscoma_pipeline/tree/developer

## Supporting information

Supplementary Figure 1: ATC categorry counts of the drugs recommended in each experiment.

## Data Availability

All data produced are available online at https://github.com/YasCoMa/caliscoma_pipeline and https://github.com/YasCoMa/screendop

https://github.com/YasCoMa/screendop

## Footnotes

1 https://portal.gdc.cancer.gov/

2 https://icgc.org/

3 https://raw.githubusercontent.com/drug2ways/results/master/networks/data/custom_network.tsv

4 https://www.ncbi.nlm.nih.gov/geo/query/acc.cgi?acc=GSE425

5 https://www.ncbi.nlm.nih.gov/geo/query/acc.cgi?acc=GSE140082

6 https://dcc.icgc.org/releases/current/Projects/LIRI-JP

7 https://dcc.icgc.org/releases/current/Projects/LIHC-US

8 https://raw.githubusercontent.com/drug2ways/results/master/validation/data/DrugBank-MeSH-slim-counts.tsv

9 https://www.cancer.gov/about-cancer/treatment/drugs/liver

10 https://go.drugbank.com/drugs/DB01169

11 https://go.drugbank.com/drugs/DB12141

## References

[1] Hua, Y., Dai, X., Xu, Y., Xing, G., Liu, H., Lu, T., Chen, Y., Zhang, Y.: Drug repositioning: Progress and challenges in drug discovery for various diseases. Eur. J. Med. Chem. 234, 114239 (2022)

[2] Li, J., Ping, Y., Li, H., Li, H., Liu, Y., Liu, B., Wang, Y.: Prognostic prediction of carcinoma by a differential-regulatory-network-embedded deep neural network. Comput. Biol. Chem. 88, 107317 (2020)

[3] Ravindran, U., Gunavathi, C.: A survey on gene expression data analysis using deep learning methods for cancer diagnosis. Prog. Biophys. Mol. Biol. 177, 1–13 (2023)

[4] Davis, K.D., Aghaeepour, N., Ahn, A.H., Angst, M.S., Borsook, D., Brenton, A., Burczynski, M.E., Crean, C., Edwards, R., Gaudilliere, B., Hergenroeder, G.W., Iadarola, M.J., Iyengar, S., Jiang, Y., Kong, J.-T., Mackey, S., Saab, C.Y., Sang, C.N., Scholz, J., Segerdahl, M., Tracey, I., Veasley, C., Wang, J., Wager, T.D., Wasan, A.D., Pelleymounter, M.A.: Discovery and validation of biomarkers to aid the development of safe and effective pain therapeutics: challenges and opportunities. Nat. Rev. Neurol. 16(7), 381–400 (2020)

[5] Olivier, M., Asmis, R., Hawkins, G.A., Howard, T.D., Cox, L.A.: The need for Multi-Omics biomarker signatures in precision medicine. Int. J. Mol. Sci. 20(19) (2019)

[6] Clough, E., Barrett, T.: The gene expression omnibus database. Methods Mol. Biol. 1418, 93–110 (2016)

[7] Huang, S., Yee, C., Ching, T., Yu, H., Garmire, L.X.: A novel model to combine clinical and pathway-based transcriptomic information for the prognosis prediction of breast cancer. PLoS Comput. Biol. 10(9), 1003851 (2014)

[8] Schupp, J.C., Vukmirovic, M., Kaminski, N., Prasse, A.: Transcriptome profiles in sarcoidosis and their potential role in disease prediction. Curr. Opin. Pulm. Med. 23(5), 487–492 (2017)

[9] Zhu, W., Xie, L., Han, J., Guo, X.: The application of deep learning in cancer prognosis prediction. Cancers 12(3) (2020)

[10] Adam, G., Rampášek, L., Safikhani, Z., Smirnov, P., Haibe-Kains, B., Goldenberg, A.: Machine learning approaches to drug response prediction: challenges and recent progress. NPJ Precis Oncol 4, 19 (2020)

[11] Vesteghem, C., Brøndum, R.F., Sønderkær, M., Sommer, M., Schmitz, A., Bødker, J.S., Dybkær, K., El-Galaly, T.C., Bøgsted, M.: Implementing the FAIR data principles in precision oncology: review of supporting initiatives. Brief. Bioinform. 21(3), 936–945 (2020)

[12] Niarakis, A., Waltemath, D., Glazier, J., Schreiber, F., Keating, S.M., Nickerson, D., Chaouiya, C., Siegel, A., Noël, V., Hermjakob, H., Helikar, T., Soliman, S., Calzone, L.: Addressing barriers in comprehensiveness, accessibility, reusability, interoperability and reproducibility of computational models in systems biology. Brief. Bioinform. 23(4) (2022)

[13] Borgwardt, K.M., Kriegel, H.-P., Vishwanathan, S.V.N., Schraudolph, N.N.: Graph kernels for disease outcome prediction from protein-protein interaction networks. Pac. Symp. Biocomput., 4–15 (2007)

[14] Golriz Khatami, S., Mubeen, S., Bharadhwaj, V.S., Kodamullil, A.T., Hofmann-Apitius, M., Domingo-Fernández, D.: Using predictive machine learning models for drug response simulation by calibrating patient-specific pathway signatures. NPJ Syst Biol Appl 7(1), 40 (2021)

[15] Das, J., Yu, H.: HINT: High-quality protein interactomes and their applications in understanding human disease. BMC Syst. Biol. 6, 92 (2012)

[16] Siglidis, G., Nikolentzos, G., Limnios, S., Giatsidis, C., Skianis, K., Vazirgiannis, M.: GraKeL: A graph kernel library in python (2018) arXiv:1806.02193 [stat.ML]

[17] Kriege, N.M., Johansson, F.D., Morris, C.: A survey on graph kernels. Applied Network Science 5(1), 1–42 (2020)

[18] Borgwardt, K., Ghisu, E., Llinares-López, F., O’Bray, L., Rieck, B.: Graph kernels: State-of-the-Art and future challenges. Foundations and Trends® in Machine Learning 13(5-6), 531–712 (2020)

[19] Cervantes, J., Garcia-Lamont, F., Rodríguez-Mazahua, L., Lopez, A.: A comprehensive survey on support vector machine classification: Applications, challenges and trends. Neurocomputing 408, 189–215 (2020)

[20] Vujović, Ž.: Classification model evaluation metrics. Journal of Advanced Computer Science and … (2021)

[21] Fang, Z., Liu, X., Peltz, G.: GSEApy: a comprehensive package for performing gene set enrichment analysis in python. Bioinformatics 39(1) (2023)

[22] Olatunji, S.O., Alansari, A., Alkhorasani, H., Alsubaii, M., Sakloua, R., Alzahrani, R., Alsaleem, Y., Alassaf, R., Farooqui, M., Basheer Ahmed, M.I., Alhiyafi, J.: Preemptive diagnosis of alzheimer’s disease in the eastern province of saudi arabia using computational intelligence techniques. Comput. Intell. Neurosci. 2022, 5476714 (2022)

[23] Elreedy, D., Atiya, A.F.: A comprehensive analysis of synthetic minority oversampling technique (SMOTE) for handling class imbalance. Inf. Sci. 505, 32–64 (2019)

[24] Chawla, N.V., Bowyer, K.W., Hall, L.O., Kegelmeyer, W.P.: SMOTE: Synthetic minority over-sampling technique. jair 16, 321–357 (2002)

[25] Kasprzyk, A.: BioMart: driving a paradigm change in biological data management. Database 2011, 049 (2011)

[26] Zmrzlikar, J., Žganec, M., Ausec, L., Štajdohar, M.: RNAnorm: RNA-seq Data Normalization in Python. https://github.com/genialis/RNAnorm

[27] Hans, C.: Elastic net regression modeling with the orthant normal prior. J. Am. Stat. Assoc. 106(496), 1383–1393 (2011)

[28] Ogutu, J.O., Schulz-Streeck, T., Piepho, H.-P.: Genomic selection using regularized linear regression models: ridge regression, lasso, elastic net and their extensions. BMC Proc. 6 **Suppl 2**(Suppl 2), 10 (2012)

[29] Akiba, T., Sano, S., Yanase, T., Ohta, T., Koyama, M.: Optuna: A nextgeneration hyperparameter optimization framework. In: Proceedings of the 25th ACM SIGKDD International Conference on Knowledge Discovery & Data Mining. KDD ’19, pp. 2623–2631. Association for Computing Machinery, New York, NY, USA (2019)

[30] Barrett, T., Suzek, T.O., Troup, D.B., Wilhite, S.E., Ngau, W.-C., Ledoux, P., Rudnev, D., Lash, A.E., Fujibuchi, W., Edgar, R.: NCBI GEO: mining millions of expression profiles–database and tools. Nucleic Acids Res. 33(Database issue), 562–6 (2005)

[31] Bullinger, L., Ehrich, M., Döhner, K., Schlenk, R.F., Döhner, H., Nelson, M.R., Boom, D.: Quantitative DNA methylation predicts survival in adult acute myeloid leukemia. Blood 115(3), 636–642 (2010)

[32] Kommoss, S., Winterhoff, B., Oberg, A.L., Konecny, G.E., Wang, C., Riska, S.M., Fan, J.-B., Maurer, M.J., April, C., Shridhar, V., Kommoss, F., Bois, A., Hilpert, F., Mahner, S., Baumann, K., Schroeder, W., Burges, A., Canzler, U., Chien, J., Embleton, A.C., Parmar, M., Kaplan, R., Perren, T., Hartmann, L.C., Goode, E.L., Dowdy, S.C., Pfisterer, J.: Bevacizumab may differentially improve ovarian cancer outcome in patients with proliferative and mesenchymal molecular subtypes. Clin. Cancer Res. 23(14), 3794–3801 (2017)

[33] UniProt Consortium: UniProt: a worldwide hub of protein knowledge. Nucleic Acids Res. 47(D1), 506–515 (2019)

[34] Love, M., Anders, S., Huber, W.: Differential analysis of count data–the DESeq2 package. Genome Biol. 15(550), 10–1186 (2014)

[35] Molania, R., Foroutan, M., Gagnon-Bartsch, J.A., Gandolfo, L.C., Jain, A., Sinha, A., Olshansky, G., Dobrovic, A., Papenfuss, A.T., Speed, T.P.: Removing unwanted variation from large-scale RNA sequencing data with PRPS. Nat. Biotechnol. 41(1), 82–95 (2023)

[36] Bullinger, L., Döhner, K., Bair, E., Fröhling, S., Schlenk, R.F., Tibshirani, R., Döhner, H., Pollack, J.R.: Use of gene-expression profiling to identify prognostic subclasses in adult acute myeloid leukemia. N. Engl. J. Med. 350(16), 1605–1616 (2004)

[37] Ritchie, M.E., Phipson, B., Wu, D., Hu, Y., Law, C.W., Shi, W., Smyth, G.K.: limma powers differential expression analyses for RNA-sequencing and microarray studies. Nucleic Acids Res. 43(7), 47 (2015)

[38] Karimizadeh, E., Sharifi-Zarchi, A., Nikaein, H., Salehi, S., Salamatian, B., Elmi, N., Gharibdoost, F., Mahmoudi, M.: Analysis of gene expression profiles and protein-protein interaction networks in multiple tissues of systemic sclerosis. BMC Med. Genomics 12(1), 1–12 (2019)

[39] Tian, L., Chen, T., Lu, J., Yan, J., Zhang, Y., Qin, P., Ding, S., Zhou, Y.: Integrated Protein-Protein interaction and weighted gene co-expression network analysis uncover three key genes in hepatoblastoma. Front Cell Dev Biol 9, 631982 (2021)

[40] Yadalam, P.K., Krishnamurthi, I., Srimathi, R., Alzahrani, K.J., Mugri, M.H., Sayed, M., Almadi, K.H., Alkahtany, M.F., Almagbol, M., Bhandi, S., Baeshen, H.A., Raj, A.T., Patil, S.: Gene and protein interaction network analysis in the epithelial-mesenchymal transition of hertwig’s epithelial root sheath reveals periodontal regenerative drug targets – an in silico study. Saudi J. Biol. Sci. 29(5), 3822–3829 (2022)

[41] Gliozzo, J., Perlasca, P., Mesiti, M., Casiraghi, E., Vallacchi, V., Vergani, E., Frasca, M., Grossi, G., Petrini, A., Re, M., Paccanaro, A., Valentini, G.: Network modeling of patients’ biomolecular profiles for clinical phenotype/outcome prediction. Sci. Rep. 10(1), 3612 (2020)

[42] Lu, H., Uddin, S.: Disease prediction using graph machine learning based on electronic health data: A review of approaches and trends. Healthc. Pap. 11(7), 1031 (2023)

[43] Li, Y., Lu, X., Zhang, J., Liu, Q., Zhou, D., Deng, X., Qiu, Y., Chen, Q., Li, M., Yang, G., Zheng, H., Dai, J.: Significance of parkinson family genes in the prognosis and treatment outcome prediction for lung adenocarcinoma. Front Mol Biosci 8, 735263 (2021)

[44] Allahyar, A., Ubels, J., Ridder, J.: A data-driven interactome of synergistic genes improves network-based cancer outcome prediction. PLoS Comput. Biol. 15(2), 1006657 (2019)

[45] Allahyar, A., Ridder, J.: FERAL: network-based classifier with application to breast cancer outcome prediction. Bioinformatics 31(12), 311–9 (2015)

[46] Bhardwaj, N., Lu, H.: Correlation between gene expression profiles and proteinprotein interactions within and across genomes. Bioinformatics 21(11), 2730–2738 (2005)

[47] Nevins, J.R., Huang, E.S., Dressman, H., Pittman, J., Huang, A.T., West, M.: Towards integrated clinico-genomic models for personalized medicine: combining gene expression signatures and clinical factors in breast cancer outcomes prediction. Hum. Mol. Genet. 12 **Spec No 2**, 153–7 (2003)

[48] Sadaf, N., Kumar, N., Ali, M., Ali, V., Bimal, S., Haque, R.: Arsenic trioxide induces apoptosis and inhibits the growth of human liver cancer cells. Life Sci. 205, 9–17 (2018)

[49] Wang, H.-Y., Zhang, B., Zhou, J.-N., Wang, D.-X., Xu, Y.-C., Zeng, Q., Jia, Y.-L., Xi, J.-F., Nan, X., He, L.-J., Yue, W., Pei, X.-T.: Arsenic trioxide inhibits liver cancer stem cells and metastasis by targeting SRF/MCM7 complex. Cell Death Dis. 10(6), 1–16 (2019)

[50] Zhang, X., Hu, B., Sun, Y.-F., Huang, X.-W., Cheng, J.-W., Huang, A., Zeng, H.-Y., Qiu, S.-J., Cao, Y., Fan, J., Zhou, J., Yang, X.-R.: Arsenic trioxide induces differentiation of cancer stem cells in hepatocellular carcinoma through inhibition of LIF/JAK1/STAT3 and NF-kB signaling pathways synergistically. Clin. Transl. Med. 11(2), 335 (2021)

[51] Rautenberg, C., Germing, U., Haas, R., Kobbe, G., Schroeder, T.: Relapse of acute myeloid leukemia after allogeneic stem cell transplantation: Prevention, detection, and treatment. Int. J. Mol. Sci. 20(1) (2019)

[52] Zhang, T., Merle, P., Wang, H., Zhao, H., Kudo, M.: Combination therapy for advanced hepatocellular carcinoma: do we see the light at the end of the tunnel? Hepatobiliary Surg. Nutr. 10(2), 180–192 (2021)

[53] Wang, L., Min, Z., Wang, X., Hu, M., Song, D., Ren, Z., Cheng, Y., Wang, Y.: Arsenic trioxide and sorafenib combination therapy for human hepatocellular carcinoma functions via up-regulation of TNF-related apoptosis-inducing ligand. Oncol. Lett. 16(3), 3341–3350 (2018)

[54] Zhu, Y., Brettin, T., Evrard, Y.A., Partin, A., Xia, F., Shukla, M., Yoo, H., Doroshow, J.H., Stevens, R.L.: Ensemble transfer learning for the prediction of anti-cancer drug response. Sci. Rep. 10(1), 18040 (2020)

