## Supplementary figures and images for "Multi-task analysis of gene expression data on cancer public datasets"

### Supplementary Figure 1: ATC categorry counts of the drugs recommended in each experiment.

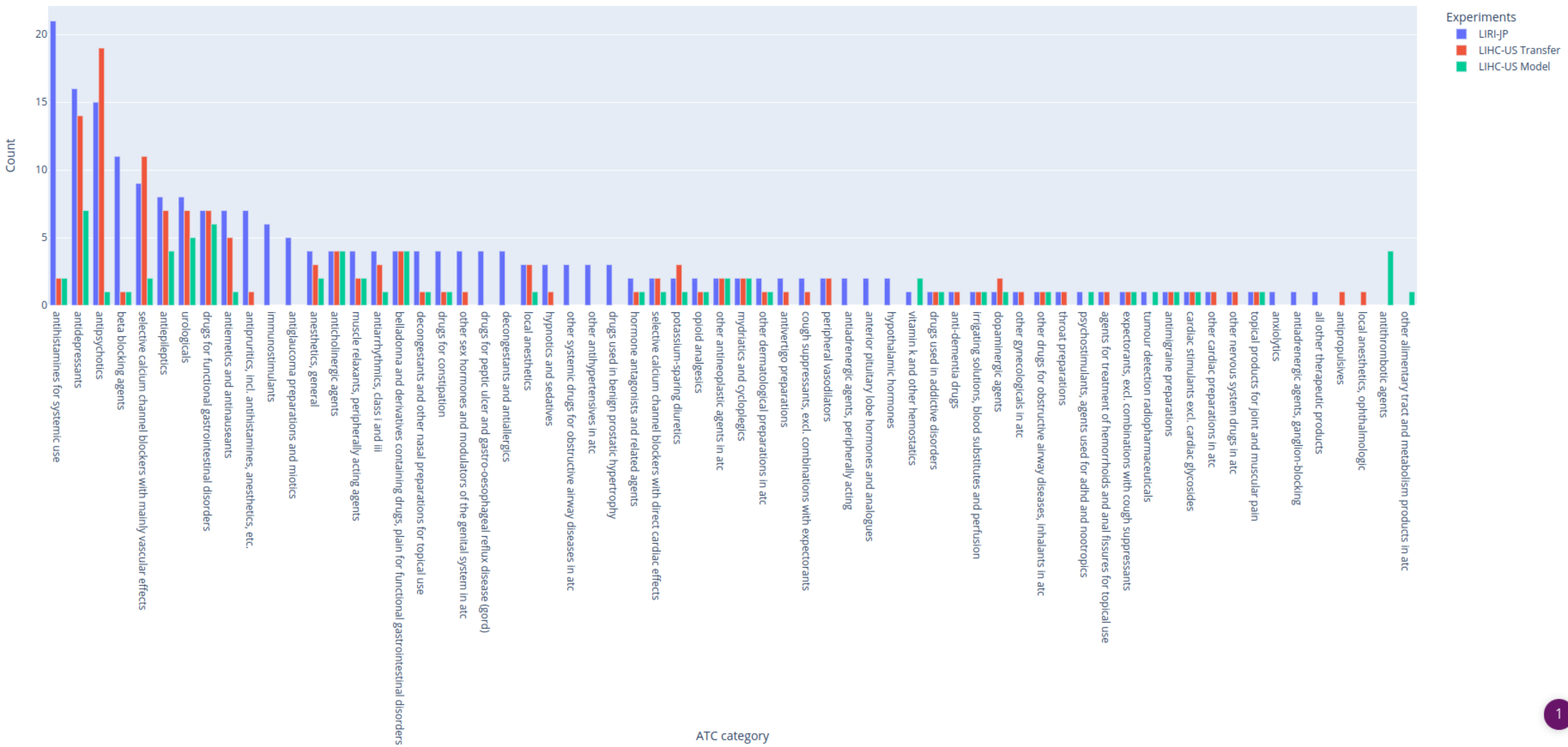
